# COVID-19 in patients undergoing renal replacement therapy in Scotland: findings and experience from the Scottish Renal Registry

**DOI:** 10.1101/2020.07.12.20148197

**Authors:** Samira Bell, Jacqueline Campbell, Jackie McDonald, Martin O’Neill, Chrissie Watters, Katharine Buck, Zoe Cousland, Mark Findlay, Nazir I Lone, Wendy Metcalfe, Shona Methven, Robert Peel, Alison Almond, Vinod Sanu, Elaine Spalding, Peter C Thomson, Patrick B Mark, Jamie P Traynor, on behalf of the Scottish Renal Registry

**Author notes:** Corresponding Author: Dr Samira Bell, Division of Population Health and Genomics, School of Medicine, University of Dundee, Dundee, DD1 9SY.

## Abstract

**Introduction:** Infection with the severe acute respiratory coronavirus 2 (SARS-CoV-2) has led to a worldwide pandemic with coronavirus disease 2019 (COVID-19), the disease caused by SARS-CoV-2, overwhelming healthcare systems globally. Preliminary reports suggest a high incidence of infection and mortality with SARS-CoV-2 in patients receiving renal replacement therapy (RRT). The aims of this study are to report characteristics, rates and outcomes of all patients affected by infection with SARS-CoV-2 undergoing RRT in Scotland.

**Methods:** Study design was an observational cohort study. Data were linked between the Scottish Renal Registry, Health Protection Scotland and the Scottish Intensive Care Society Audit Group national data sets using a unique patient identifier (Community Health Index (CHI)) for each individual by the Public Health and Intelligence unit of Public Health, Scotland. Descriptive statistics and survival analyses were performed.

**Results:** During the period 1^st^ March 2020 to 31^st^ May 2020, 110 patients receiving RRT tested positive for SARS-CoV-2 amounting to 2% of the prevalent RRT population. Of those affected, 87 were receiving haemodialysis or peritoneal dialysis and 24 had a renal transplant. Patients who tested positive were older and more likely to reside in more deprived postcodes. Mortality was high at 26.7% in the dialysis patients and 29.2% in the transplant patients.

**Conclusion:** The rate of detected SARS-CoV-2 in people receiving RRT in Scotland was relatively low but with a high mortality for those demonstrating infection. Although impossible to confirm, it appears that the measures taken within dialysis units coupled with the national shielding policy, have been effective in protecting this population from infection.

## Introduction

Infection with the severe acute respiratory coronavirus 2 (SARS-CoV-2) has led to a worldwide pandemic with COVID-19, the disease caused by SARS-CoV-2, overwhelming healthcare systems globally ^1^. The clinical course of the disease varies significantly from asymptomatic disease to multi-organ failure and death^2^. Patients receiving renal replacement therapy (RRT) are a vulnerable population as those receiving dialysis are usually older with significant co-morbidity and require regular attendance at a healthcare facility which further increases their risk of exposure. Patients who have a kidney transplant are presumed to be at high risk due to their obligate immunosuppressive medications. There have been a number of published recommendations on how to mitigate spread of SARS-CoV-2 in dialysis units ^3-7^. There is, however, a lack of data regarding clinical course and outcome in patients receiving RRT following infection with SARS-CoV-2. Furthermore, early reports suggest that case fatality in dialysis patients if infected is high with reports suggesting fatality in the order of 5-20% compared to 1-2% in patients not requiring RRT ^8-10^. Patients requiring RRT often have multiple comorbid conditions which are widely documented as increasing the risk of death from COVID-19 such as older age, chronic conditions such as diabetes mellitus and hypertension ^11,12^ but may be at additional risk due to the relatively impaired immune response associated with kidney failure.

The first case of SARS-CoV-2 infection in Scotland was confirmed on 1^st^ March 2020, although subsequent data suggest that the virus had been in circulation for weeks prior to this ^13^. On 24^th^ March, as part of the Scottish Government response to the pandemic, certain groups of patients were identified as the most vulnerable to developing severe illness and advised to “shield” meaning they were advised to remain at home and minimise all non-essential contact with other members of their household. This “shielded” category included renal transplant recipients from the outset but Scottish (and United Kingdom) government advice was not applied to for dialysis patients until 28^th^ April 2020. This posed significant challenges to renal units across the country in delivering care to these patients whilst minimising risk of exposure and preventing transmission of infection to these vulnerable patient groups.

The aims of this study are to report characteristics, rates and outcomes of all patients affected by infection with SARS-CoV-2 undergoing RRT in Scotland.

## Methods

### Data Sources

Study design was an observation cohort study. The following national data sets were linked using a unique patient identifier (Community Health Index (CHI)) for each individual by Public Health and Intelligence unit of Public Health, Scotland. The Scottish Renal Registry is a national registry of all patients receiving RRT for end stage renal disease (ESRD) in Scotland (haemodialysis, peritoneal dialysis and transplant). It was established in 1991 with data backfilled to 1960 from European Renal Association-European Dialysis and Transplant Association (ERA-EDTA) with the first patient dialysed for ESRD in Scotland in 1960. The Scottish Renal Registry has 100% unit and patient coverage. Data held by the registry include patient demographics including historical postcodes (for calculating Scottish Index of Multiple Deprivation), full RRT history (for ESRD), date and cause of death (using ERA-EDTA codes), primary renal diagnosis (using ERA-EDTA codes) and monthly linkage with National Health Service Blood and Transplant (NHS BT) for transplant status. The Scottish Government provide online calculators allowing use of patient postcode to generate divisions of socioeconomic deprivation, the Scottish Index of Multiple Deprivation (SIMD)^14^. Using patient postcode, deprivation quintiles were calculated and categorized into most deprived quintiles with 1 corresponding to most deprived and 5 as least deprived. Data on testing for COVID-19 were obtained from Health Protection Scotland with date of test and result reported. Data on admission to intensive care units were obtained from the Scottish Intensive Care Society Audit Group (SICSAG). The Scottish Intensive Care Society Audit Group (SICSAG) has maintained a national database of patients admitted to adult general Intensive Care Units (ICU) in Scotland since 1995^15^. Detailed information is produced on the management of critically ill or injured patients. All general intensive care units (ICU) and combined ICU/high dependency units (HDU) collect data. During this period, there was no limitation of access to ICU admission for patients with comorbid conditions, with patients admitted to ICU if clinicians felt it was clinically appropriate.

Testing for SARS-CoV-2 was carried out by real-time polymerase chain reaction (RT-PCR) on a combined nasal and pharyngeal throat swab only in symptomatic patients or endotracheal aspirate for those ventilated in ICU. If a patient has been tested multiple times, the date of the first positive or first negative test was used. If a patient has tested positive, then subsequently tested negative they would only appear in the positive cohort.

### Statistical Analysis

Data were linked and analysed by a Public Health Intelligence analyst at Public Health Scotland. Baseline characteristics were displayed as mean and standard deviation (SD) or median and interquartile range (IQR) for continuous variables depending on normality. T-tests and chi-squared tests were utilised to examine differences in parameters between those who died and those were still alive as of 31^st^ May 2020. Categorical variables were displayed as number and percentage. Logistic regression was performed with SARS-Cov-2 positivity as a binary outcome. Crude survival was measured at 30 days since first SARS-Cov-2 test using Kaplan Meier survival analyses. Cox proportional hazards models were performed to examine predictors of death. All analyses were performed using R Studio 3.5.1^16^.

## Results

### Scottish Renal Registry

The Scottish renal registry collates data from all 9 adult renal units in Scotland and 28 satellite haemodialysis units (Figure 1) serving a population of 5.4 million. On 31st May 2020, there were 5475 prevalent patients in Scotland receiving RRT. Of these, 1983 were on haemodialysis and 206 peritoneal dialysis and 3286 with functioning renal transplants.

**Figure 1:**
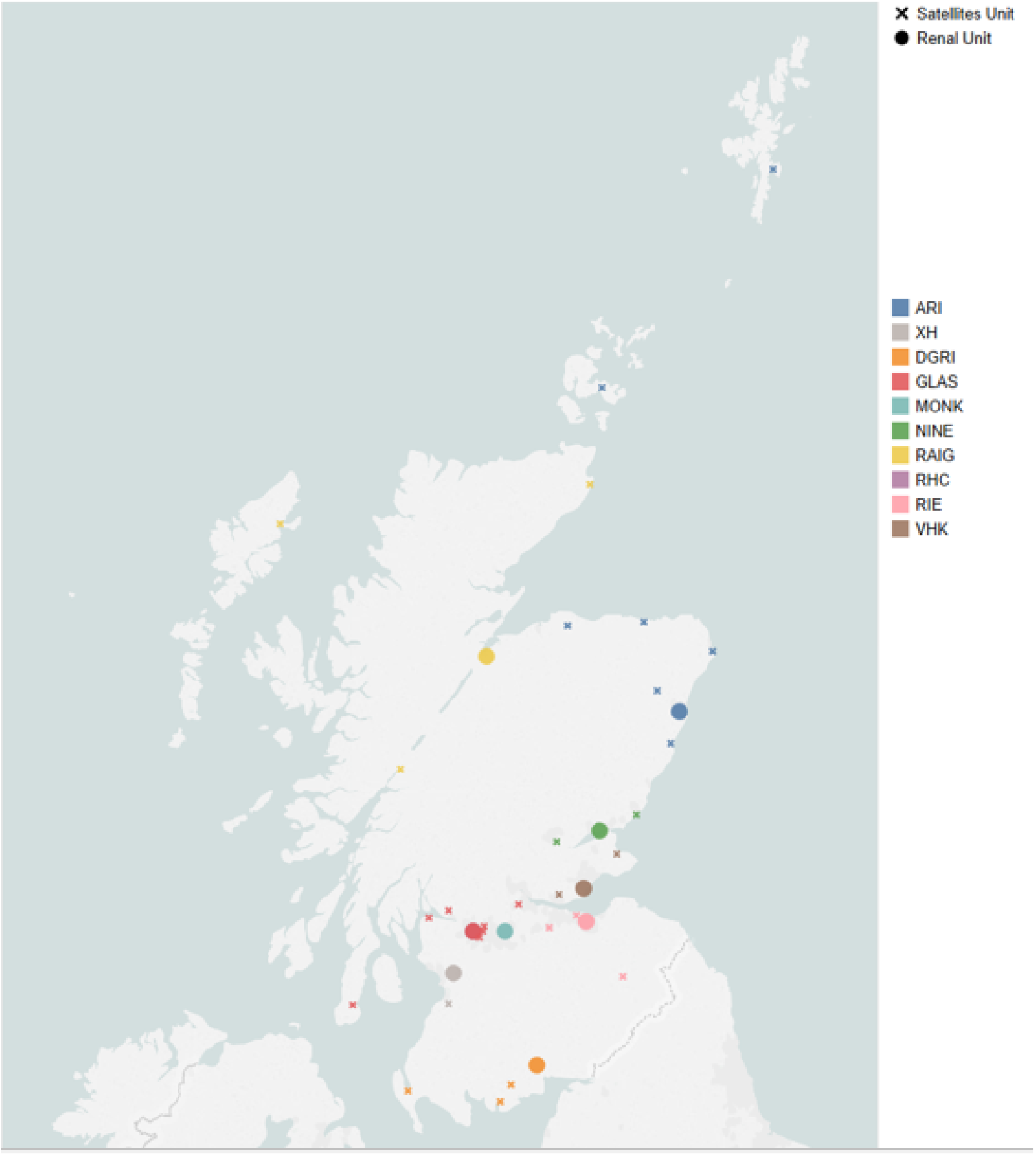
Dialysis Locations in Scotland. ARI Aberdeen Royal Infirmary, XH Crosshouse Hospital, DGRI Dumfries and Galloway Royal Infirmary, GLAS Glasgow Royal Infirmary, MONK Monklands Hospital, NINE Ninewells Hospital Dundee, RAIG Raigmore Hospital Inverness, RIE Royal Infirmary of Edinburgh, VHK Victoria Hospital Kirkcaldy.

### Characteristics of SARS-CoV-2 positive patients

As of 31^st^ May 2020, 876 patients receiving RRT had been tested for SARS-CoV-2 infection. This represents 16% of the total number of 5475 receiving RRT on 31^st^ May 2020. One hundred and ten were positive which amounts to 2% of the prevalent RRT population. Of those affected, 87 were receiving haemodialysis or peritoneal dialysis and 24 had a renal transplant. Figure 2 shows the distribution of those individuals who tested positive for SARS-CoV-2 by geographical location relative to the whole RRT population. In keeping with the national figures related to the general population, the majority of positive tests were within the central belt of Scotland. Figure 3a shows the number of positive cases by week with a peak in positive cases between the 30^th^ of March and 13^th^ of April. This contrasts with data for general population across Scotland where the number of new cases only started to fall during week beginning 27^th^ April (Figure 3b). There were 986 tests performed in 876 patients until the 31^st^ May with 110 (11.2%) testing positive. The cumulative number tested and the proportion of positive cases over time is shown in Figure 4a with a sharp increase in testing over the time period examined. Characteristics of the renal replacement population and those who tested positive for SARS-CoV-2 are shown in Table 1. Mean age of those tested positive was 64.3 years old (SD 14.4) with a primary renal diagnosis of diabetes in 33.7% and median dialysis vintage of 4 years (IQR 1-8 years). Of the 24 patients who had a functioning transplant when tested positive for SARS-CoV-2, the majority of the transplants (n=14) were performed over 10 years ago with 8% (n=2) cases recorded had a transplant less than 2 years ago. 65% of those who tested positive lived in the most deprived areas (SIMD 1 or 2). 98 of 110 who tested positive were of white ethnicity. We do not have data on the ethnicity of the entire RRT population. Using logistic regression, increasing age at time of infection OR 1.038 (95%CI 1.023-1.053), p< 0.001 and social deprivation OR 0.778 (95% CI 0.673-0.899), p=0.001 (with every quintile increase in in SIMD) were predictors of testing positive for all those on RRT on 31 May 2020 registered on the SRR.

**Table 1:**
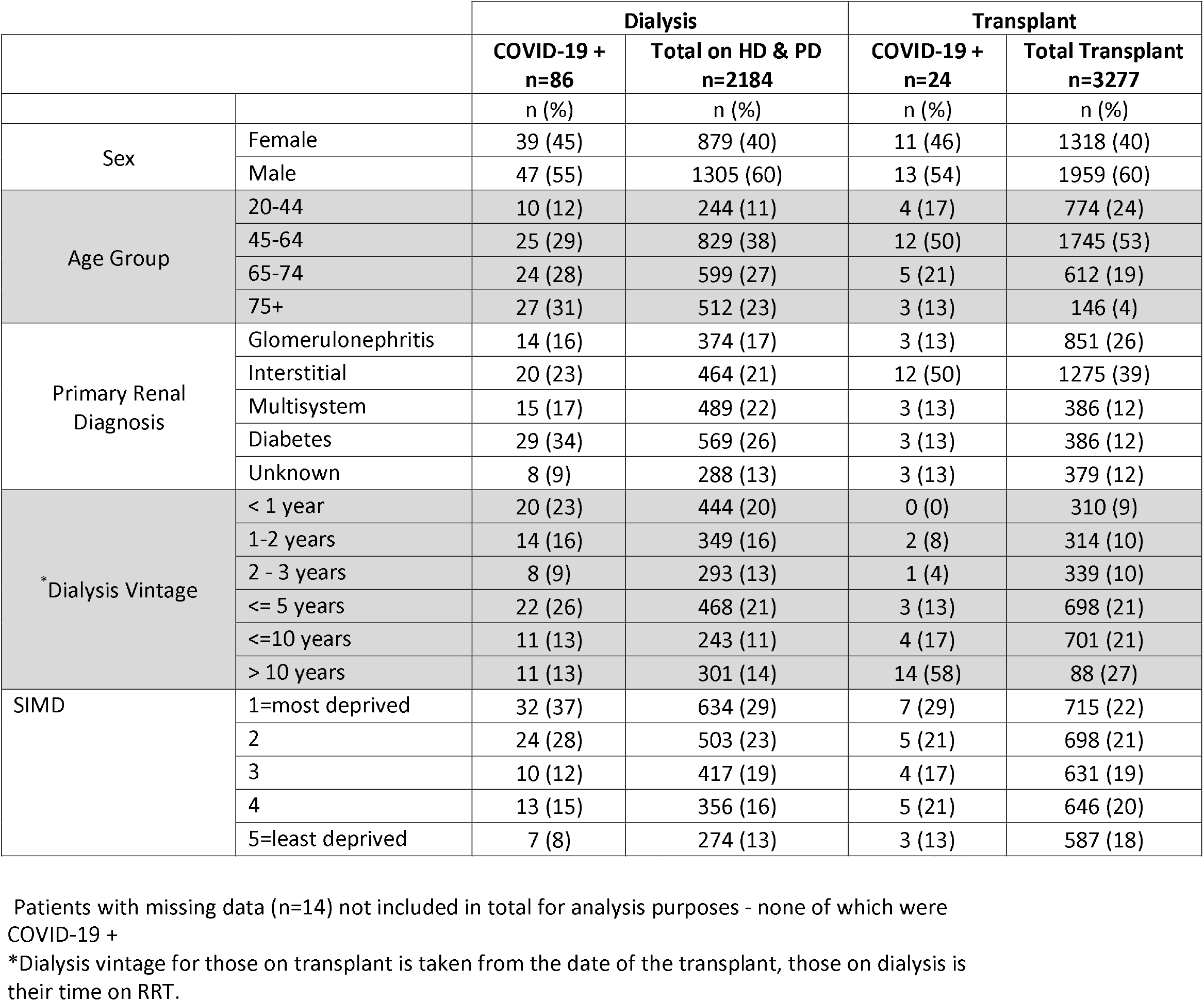
Descriptive characteristics of all prevalent patients undergoing RRT in Scotland on 31 May 2020.

**Figure 2:**
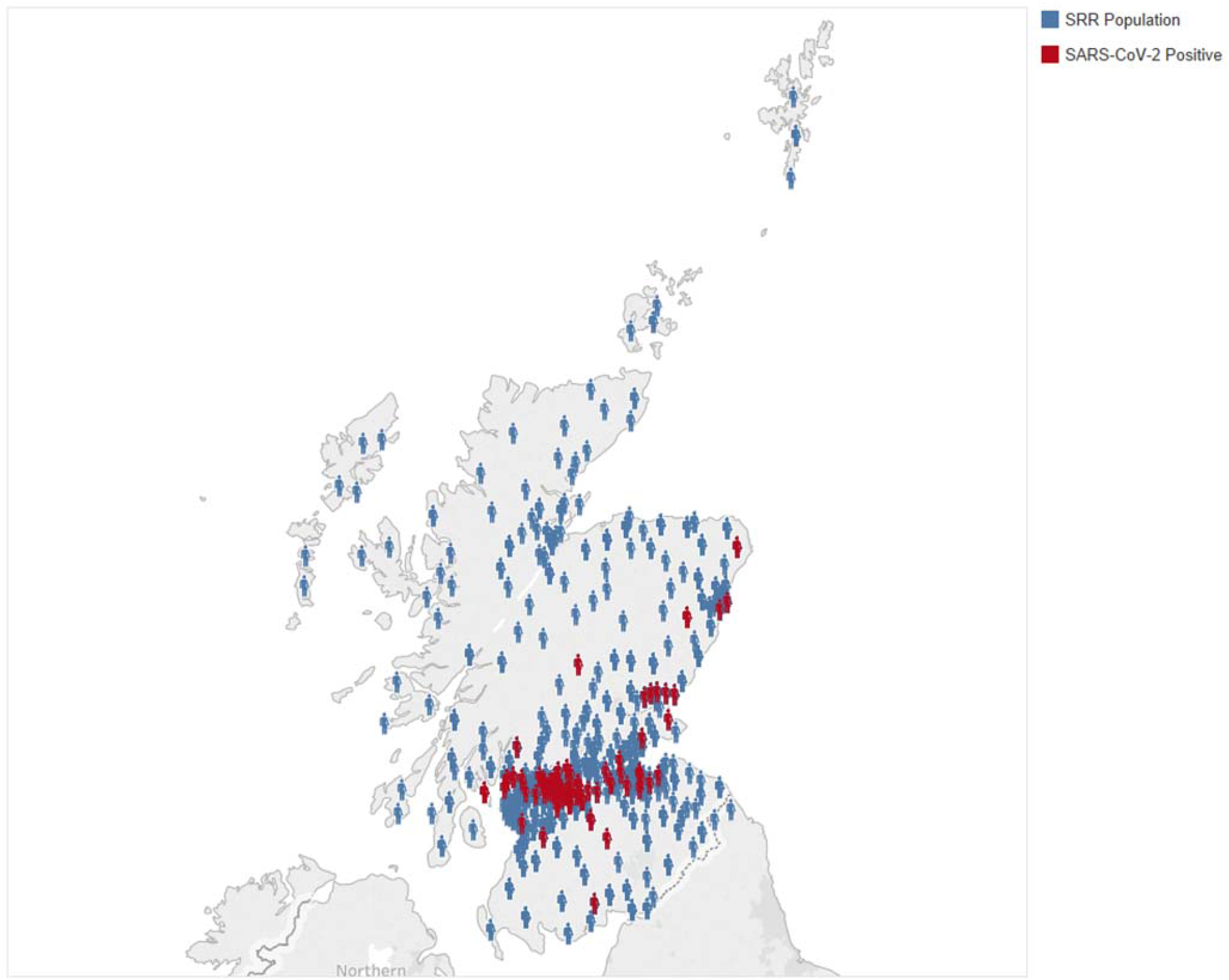
Geographical output of SARS-CoV-2 in RRT patients up to 31 May 2020.

**Figure 3a:**
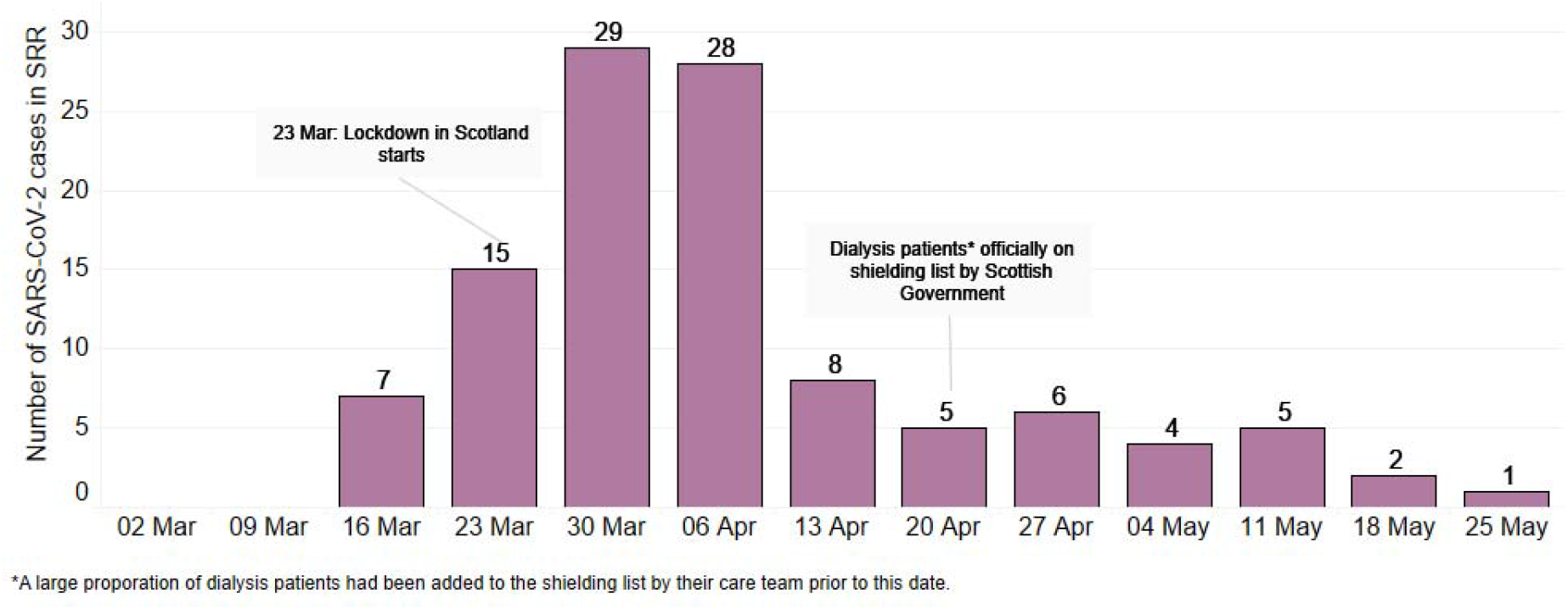
Number of patients* on RRT who tested positive for SARS-CoV-2. *If a patient has been tested multiple times, the date of the first positive or first negative test was used. If a patient has tested positive, then subsequently tested negative they would only appear in the positive cohort.

**Figure 3b:**
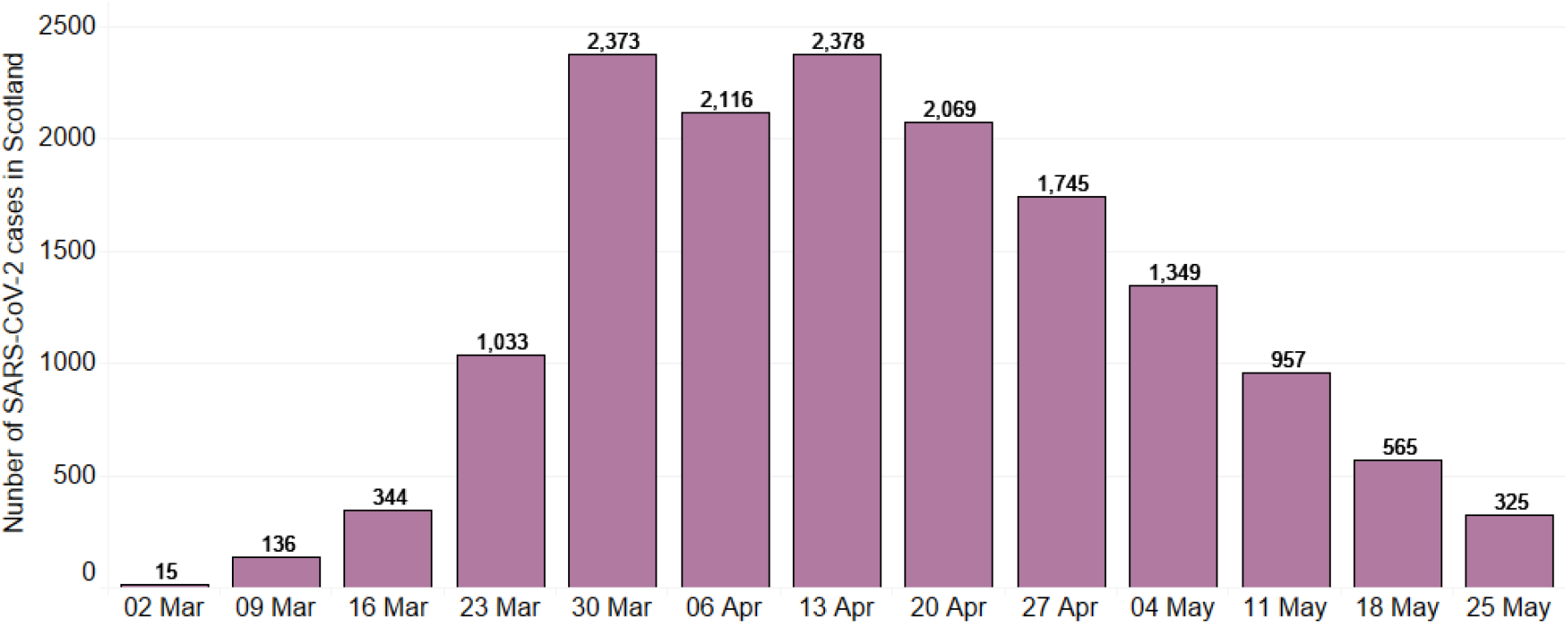
Number of patients in Scotland who tested positive for SARS-CoV-2.

**Figure 4a:**
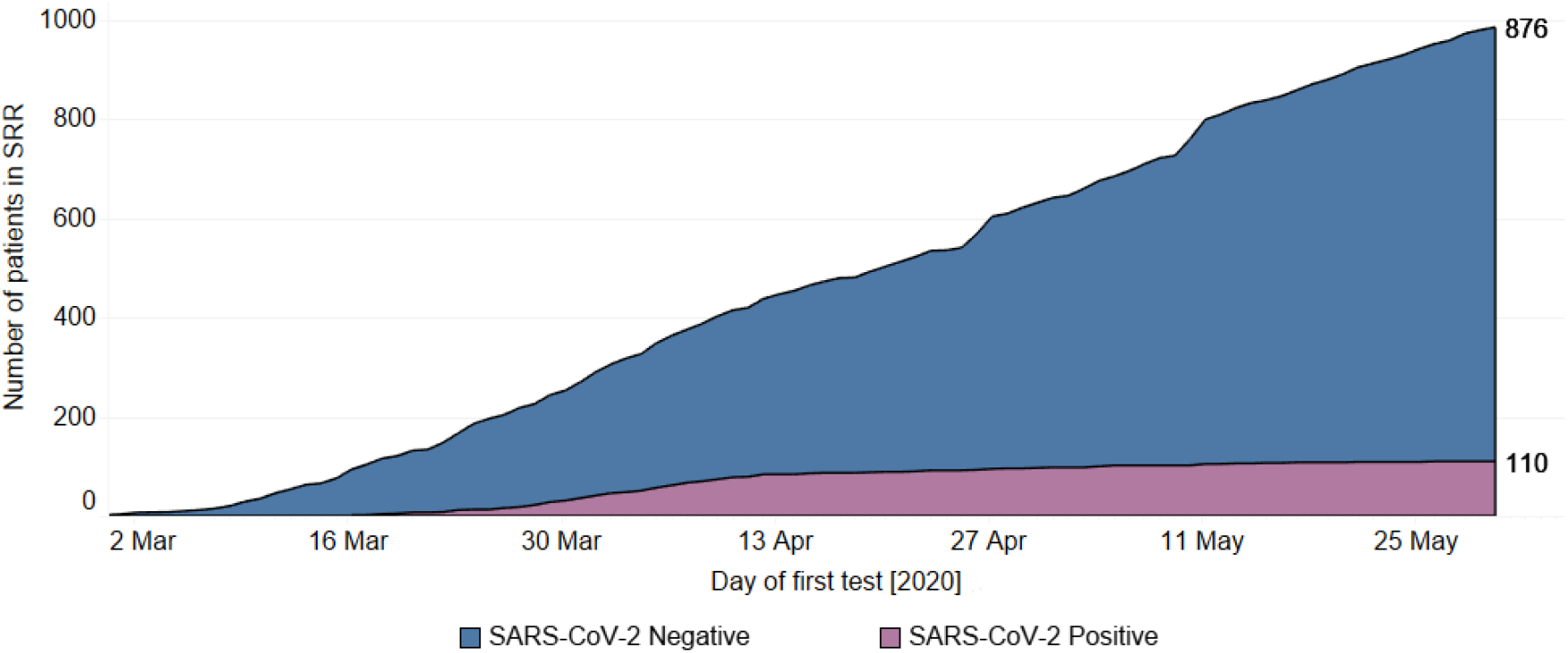
Cumulative number of SRR patients* on RRT with a SARS-CoV-2 test.

**Figure 4b:**
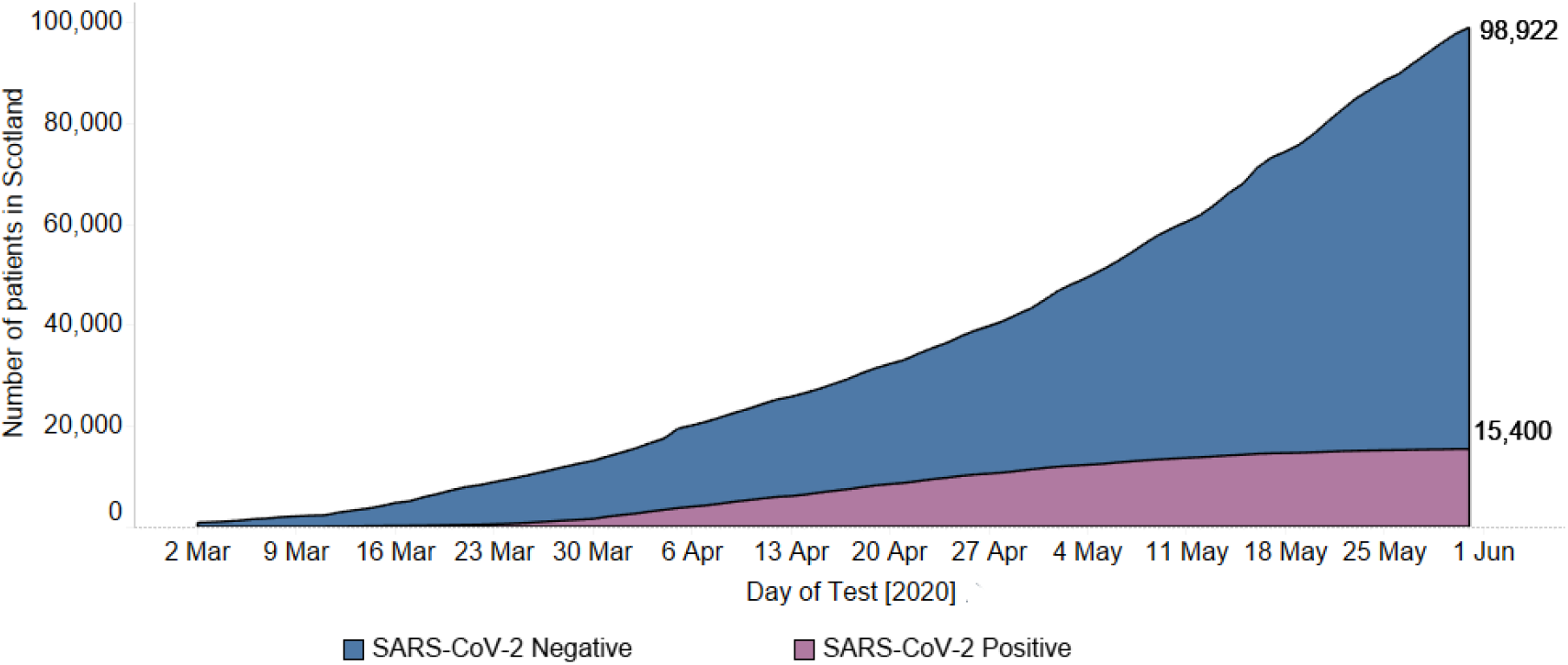
Cumulative number of patients in Scotland with a SARS-CoV-2 test.

### Patient Outcomes

As of the 31^st^ May 2020, 30 (28.2%) of the patients who tested positive for SARS-CoV-2 infection died. Twenty three (26.7%) of the 86 patients treated with haemodialysis or peritoneal dialysis who tested positive died. Seven (29.2%) of the 24 transplant patients who tested positive died. Mean age of death was higher in those who died compared with those who were still alive as of 31^st^ May 2020 (69 years (SD 11.6) vs. 62 years (SD 8.5), p=0.0182). There was no difference in median dialysis vintage of those who died compared with those who were still alive as of 31^st^ May 2020: 4 years (IQR 1-12 years) vs 4 years (IQR 1-7 years, p=0.34). One third of patients who died had a primary renal diagnosis of diabetic nephropathy. Figure 5 shows the total number of deaths in the RRT therapy population in Scotland from 1^st^ January 2020 to 31^st^ May 2020 with those who had COVID-19 identified as a primary cause of death on the death certificate. Figure 6 shows the number of deaths from 1^st^ January to 31^st^ May each year from 2017 to 2020. Deaths in 2020 not related to COVID-19 are also shown. Both figures 5 and 6 demonstrate an increase in deaths occurring in April compared with other months. Figure 5 in particular highlights that a large proportion (35%) of the deaths occurring in April were related to COVID-19. Crude thirty day survival is shown in Figure 7. There was 91% (95% CI 89-94) 30 day survival in those to tested negative compared with 71% (95% CI 63 -80) 30 day survival in those who tested positive. Increasing age was associated with mortality in cox proportional hazards model with an increase of hazard ratios for death of 1.028 (95% CI 1.001 to 1.055, p=0.038) for every year of age.

**Figure 5:**
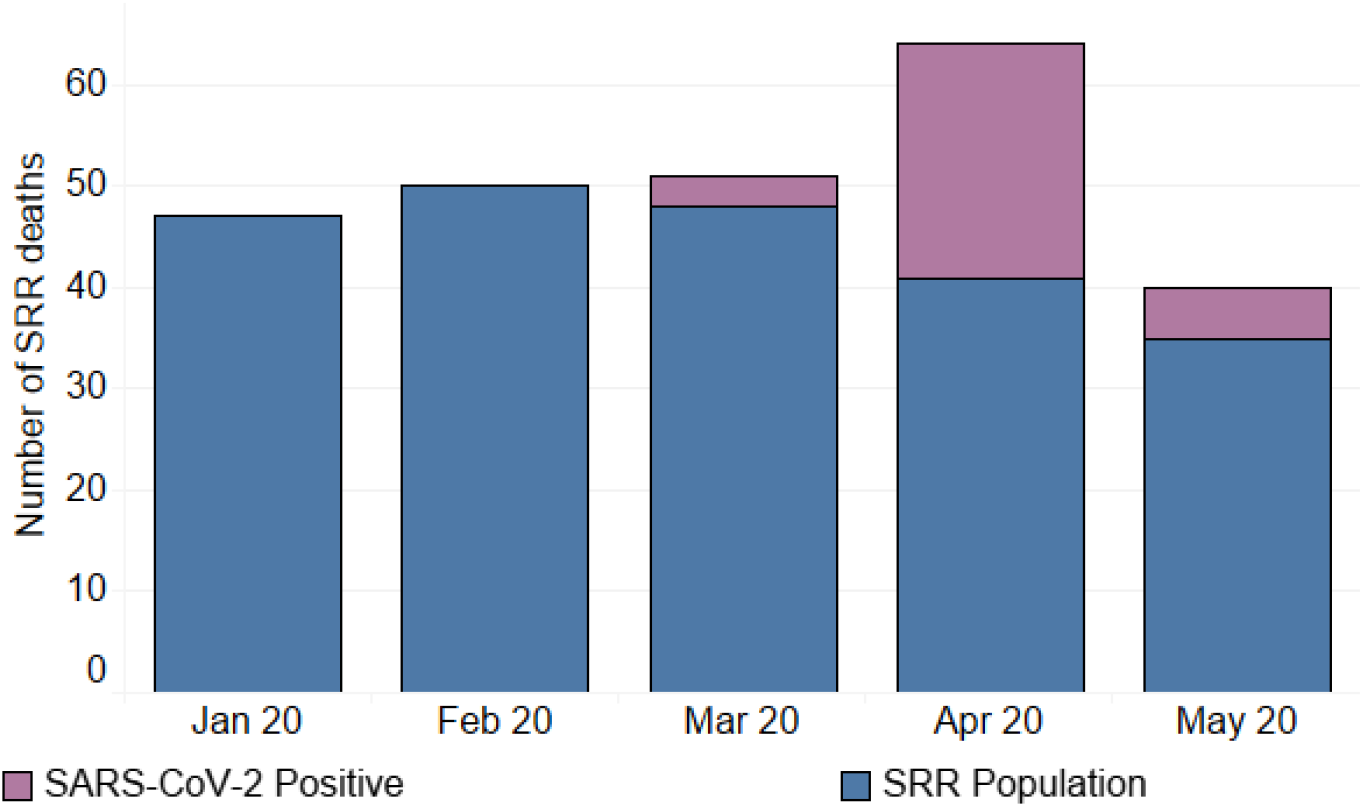
Number of Deaths in RRT population per calendar month. *For all those patients who tested positive for SARS-CoV-2 – all had SARS-CoV-2 as Cause of Death on death certificate.

**Figure 6:**
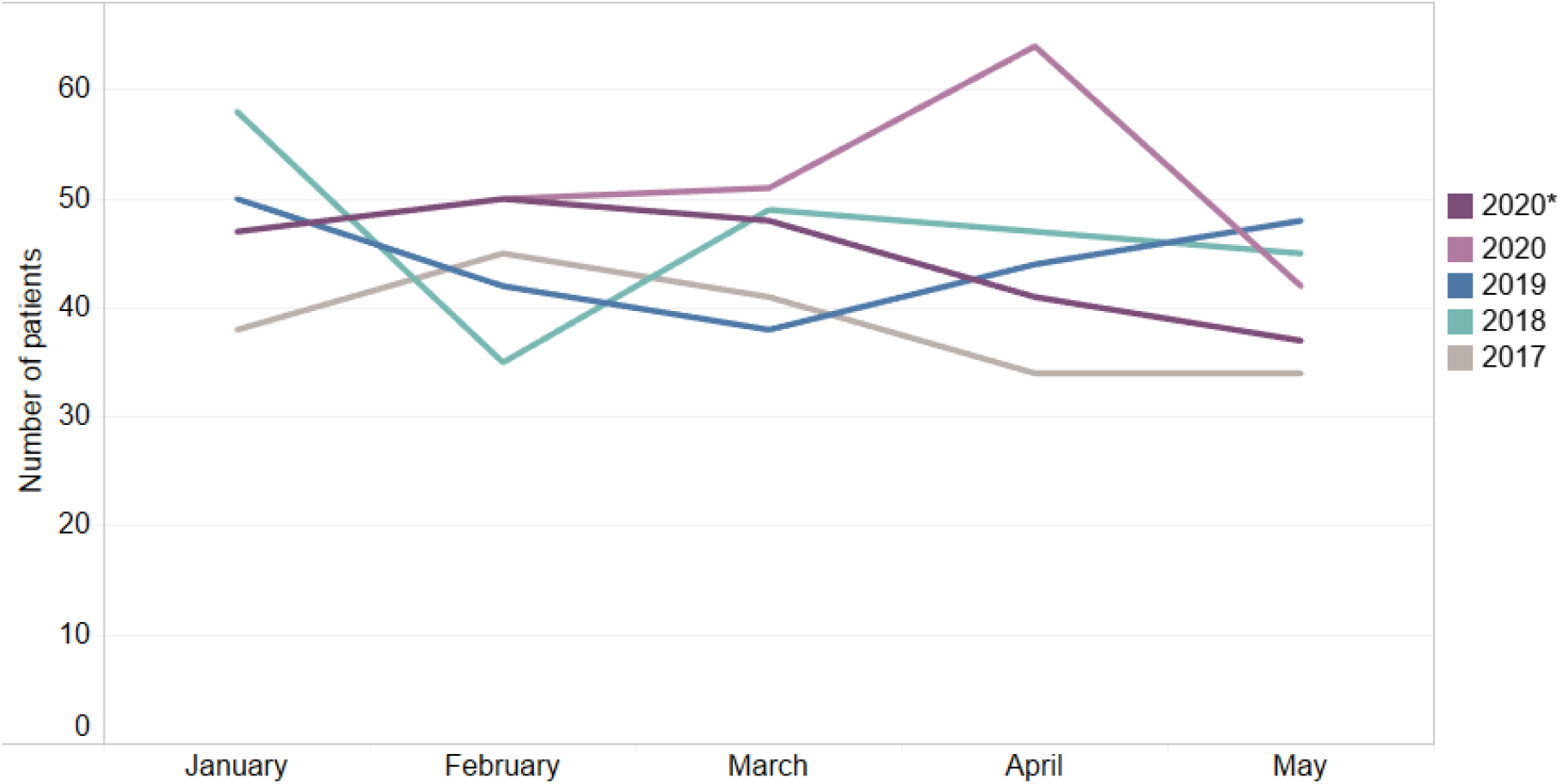
Number of deaths in RRT population – comparison over multiple years. *This lines shows the number of deaths after removing those cases where SARS-CoV-2 was the Cause of Death.

**Figure 7:**
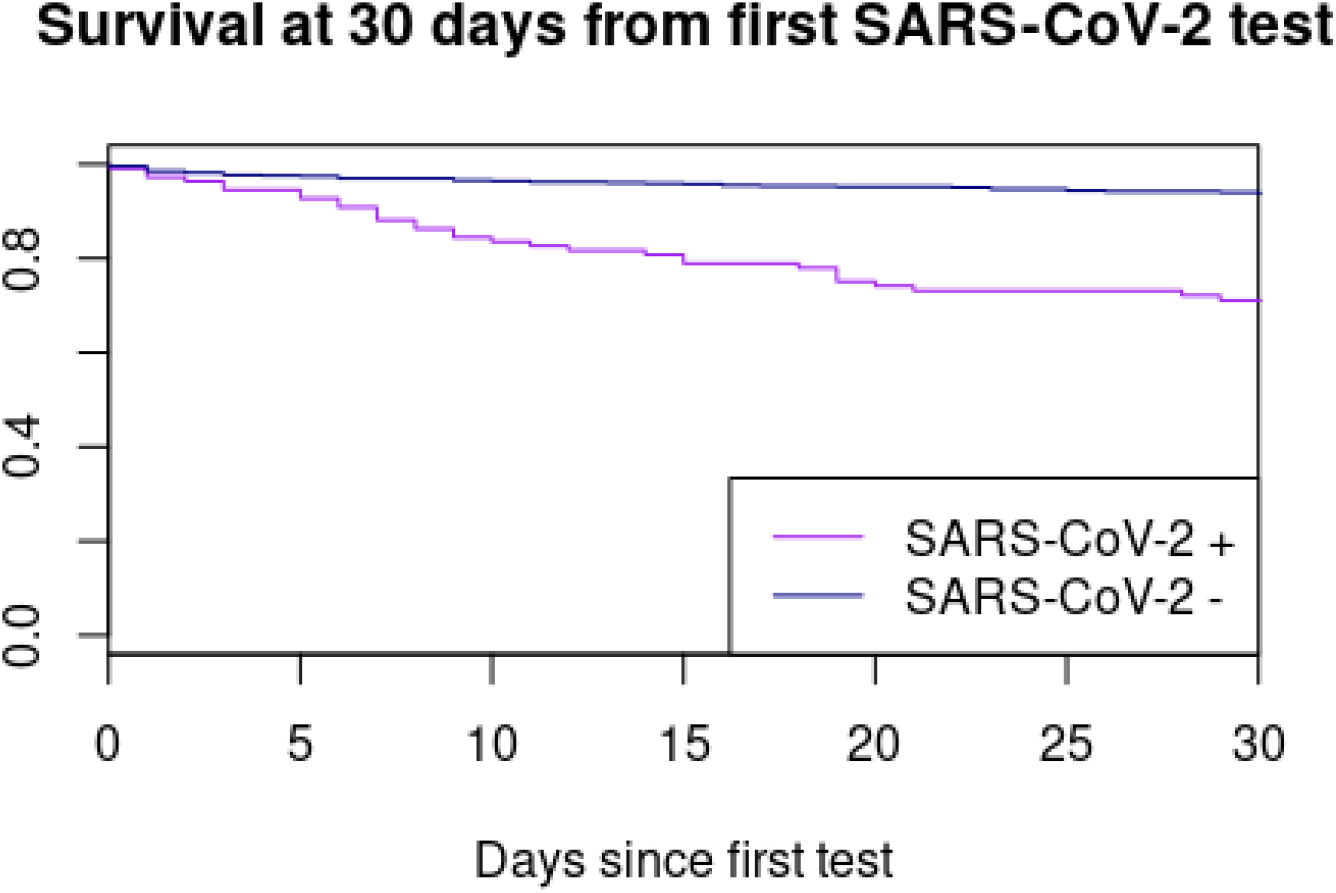
Survival at 30 days from first SARS-CoV-2 test. *Log Rank Test for KM is p<0.001

### Intensive Care Admission

During the period from 1st March 2020 to 31st May 2020 there were a total of 118 patients registered on the SRR who were admitted to an ICU/HDU in Scotland. Of these 13 (13%) patients had tested positive for SARS-CoV-2 infection. 92.3% of those with SARS-CoV-2 were discharged alive from ICU compared with 94.1% who were not tested/tested negative. However, of those who were discharged alive from ICU/HDU 41.6% of those with a positive SARS-CoV-2 test died within 7 days of being discharged compared to 6.94% of those who did not have a positive test result.

## Discussion

As of May 31^st^ 2020, SARS-CoV-2 was detected in 2% of the prevalent Scottish RRT population (3.9% of the prevalent dialysis population and 0.7% of the prevalent transplant population). Mortality was high at 26.7% in the dialysis patients and 29.2% in the transplant patients. Patients who tested positive were more likely to be older and from more deprived postcodes. Increasing age was associated with mortality.

During the early stages of the pandemic, advice about personal protective equipment (PPE) was subject to a number of changes mainly in response to guidance issued by UK government for use of PPE in the healthcare environment. On 2^nd^ April 2020, the UK and Scottish governments issued new PPE guidelines for patients and staff in secondary care and these in turn informed the UK Renal Association recommendations for PPE in dialysis units ^7,17^. The guidance has largely remained unchanged since and essentially advocates that staff use face masks (one per session) and single use aprons and gloves with all patients, including those where COVID was not suspected. After discussion with local microbiology/infectious disease/infection control teams, most units implemented these changes in renal dialysis units very quickly with all units following this guidance by 6^th^ April 2020. Two units adopted this approach before the government advice with the earliest adoption of PPE on 13th March 2020.

All dialysis patients now wear a mask for travelling into dialysis and during the session. Patients are given a mask to use for next session. Patients are advised to minimise time spent eating or drinking i.e. time with mask off. Waiting areas are a potential challenge as in some units these are very small and cannot facilitate adequate social distancing. Approaches to address this includes staggering start times for dialysis and asking patients to wait in their transport vehicle until the staff are ready to commence their dialysis session. All patients have individual transport except a few cases where following an adequate risk assessment, two patients share an ambulance but wear masks. Individualised transport has only been possible whilst there is reduced demand from other specialties which have put non-essential services on hold or at significant cost to the units by providing single occupancy taxi for transport. Most units have, or are actively making plans for, the time when individual transport is no longer available or manageable. All units measure patient temperature on arrival. The threshold for action varies between temperature of 37.8° and 38.0° degrees with the exception of one satellite unit which has a threshold of 37.0°. Once reached, this threshold triggers testing for SARS-COV2 and dialysis in isolation. All units have taken measures to minimise visits to hospital for people with a functioning transplant. Clinical records are reviewed in advance of clinics. Unless a face-to-face consultation is considered necessary, alternative options have included telephone consultations, blood tests only and increased time between clinics. Phone consultations are provided mostly by medical staff but some units include transplant nurse specialists. Bloods have been taken in a variety of settings including newly set up ‘bloods only’ clinics, bespoke mobile phlebotomy service, existing community hubs or in coordination with primary care.

There is currently a lack of data on how SARS-CoV-2 affects patients receiving haemodialysis. Early reports from Wuhan suggested a milder disease in these patients ^18^. However, more recent reports are suggesting worse outcomes. The Lombardy and Brescia regions of Italy were severely affected by COVID-19. An observational study from 4 dialysis centres in the Brescia region of Italy showed that 95 (15%) of 643 haemodialysis patients tested positive for COVID-19 with 27 (29%) of these dying ^19^. A further observational study from a haemodialysis centre in Spain found that 36 patients out of 282 patients tested positive for COVID-19 and 11 (30.5%) of these patients subsequently died. Dialysis vintage was associated with mortality in this cohort of patients ^20^. Similarly, a study of 59 consecutive patients testing positive from Columbia University in the United States found a mortality rate of 31% (18 patients)^21^. A report from the COVID-19 registry of the Spanish Society of Nephrology reported a mortality of 23% in 868 patients with SARS-CoV-2 infection. Mortality was associated with increasing age in this cohort^22^. A study from a single urban centre in the United Kingdom showed much higher rates of transmission with over 20% of their haemodialysis patients affected within 6 weeks of detection of their first case ^23^. Similar mortality rates was demonstrated in this series with 20.3% patients dying during follow up, again with increasing age being associated with higher likelihood of mortality. The outcomes we report in kidney transplant recipients with 29.2% of those with COVID-19 dying is similar to that reported in New York^24^.

In the general population, a number of comorbid conditions such as diabetes, hypertension and obesity have consistently been linked to increased risk of death with COVID-19 ^11^. In patients requiring RRT, it is clear that the risk attributable to requirement for RRT is such a strong risk factor for death that it is challenging to identify subgroups of dialysis or renal transplant recipients at highest risk.

SARS-CoV-2 has affected a relatively small proportion of the RRT patients in Scotland. However, mortality was high in those affected and COVID-19 has been associated with excess mortality in the RRT population of Scotland during this period compared to equivalent periods during previous years. Renal unit staff and transport services have worked hard to mitigate and contain transmission within the individual dialysis and satellite units. The subsequent drop in new cases by the third week of April suggests that these measures coupled with shielding were effective. It is not possible to say with absolute certainty but the experience in a large dialysis unit in London has led to similar conclusions ^23^. Infection control measures are likely to have been beneficial in reducing transmission associated with dialysis treatment, either related to transport with other patients, or during treatment itself. However, it is not possible to dissociate the reduction in number of cases of SARS-CoV-2 in the RRT population from the fall observed in the general population in response to measures encompassed by ‘lockdown’ including closure of schools and non-essential shops, increased hand hygiene and social distancing. However, as the incidence of new SARS-CoV-2 infections specifically in patients on RRT fell prior to any fall in the incidence of SARS-CoV-2 in Scotland (Figure 3), it is plausible that the combination of measures specific to this high risk group had efficacy in driving down new infections, rather than to simply assume that falling infection in RRT patients reflected reducing community transmission of SARS-CoV-2. It is worth noting that highest numbers of SARS-CoV-2 infection was observed in patients residing in areas of highest socioeconomic deprivation. Although the numbers of infections do not necessarily track to outcomes, most UK and Scotland data demonstrate that infection rates and deaths with SARS-CoV-2 are higher in areas of socioeconomic deprivation ^25^.

A strength of our study is that we have comprehensive coverage of all patients receiving RRT in Scotland with linkage to Health Protection Scotland which holds data on all testing for SARS-CoV-2 performed in Scotland. However, limitations of our study are that we did not have data on hospitalisations, co-morbidities or smoking status. Fortunately, our numbers were small. However, this limits our ability to draw conclusions from our analyses. Furthermore we do not hold data on medication use so cannot infer whether any medications, such as renin angiotensin system inhibition, was associated with different risks of infection or adverse outcome as has been investigated elsewhere ^26,27^. Only symptomatic patients were tested, so we cannot comment on the asymptomatic rate of SARS-CoV-2 infection, this may have led to lower case ascertainment in the population of patients with kidney transplants. However, unlike in the wider UK population, testing was performed liberally, and to the best of our knowledge in all symptomatic patients requiring dialysis and in those without symptoms but who presented with fever.

In summary, we demonstrate that 2% of the RRT population had a positive test for SARS-CoV-2 during the period of study. The incidence of positive tests fell following instigation of infection control measures. Mortality is high and it is unclear if any of the therapies demonstrated to improve outcomes in patients with COVID-19 such as dexamethasone or remdesivir will have efficacy in patients requiring RRT. Patients requiring dialysis were excluded from recent remdesivir trials ^28^. It is likely that in the absence of specific therapy, and high mortality, heightened vigilance of the risk of SARS-CoV-2 infection, infection control measures including use of PPE for both staff and patients, will be required for the long term to mitigate the risk of SARS-CoV-2 in this vulnerable patient group. This may come at a cost of increasing social isolation and more limited social interaction between other patients, health care workers and wider society. Moreover, the economic cost of providing additional transport for individual patients travelling to dialysis, more dedicated nursing staff and segregated dialysis bays, as well as for additional PPE will add substantially to the existing financial burden of RRT to healthcare systems.

## Data Availability

These data are held within Public Health Intelligence, Scotland are not freely available.

## Acknowledgements

We thank Kirsty Mangin at Health Protection Scotland and Lorraine Donaldson from SICSAG for their assistance with linkage of the datasets. We thank all staff in critical care units who submitted data to the SICSAG database.

